# Metformin Is Associated with Favorable Outcomes in Patients with COVID-19 and Type 2 Diabetes Mellitus

**DOI:** 10.1101/2021.05.20.21257490

**Authors:** Zhiyuan Ma, Nishit Patel, Pranathi Vemparala, Mahesh Krishnamurthy

## Abstract

**Aim:** Coronavirus disease 2019 (COVID-19) is a new pandemic the entire world is facing since December of 2019. In this retrospective study, we aim to address whether metformin use offers a low mortality on patients with COVID-19 and pre-existing type 2 diabetes mellitus (T2DM).

**Materials and Methods:** A cohort of 1356 hospitalized patients with COVID-19 and pre-existing T2DM was analyzed by multivariate regression. Covariates that potentially confound the association were further adjusted using propensity score matching or inverse probability of treatment weighting. A meta-analysis of the present study along with 5 published studies with metformin use prior to admission in patients with COVID-19 and T2DM was carried out.

**Results:** We found that metformin therapy prior to admission in patients with COVID-19 and T2DM was significantly associated with less primary outcome events including in-hospital mortality and hospice care enrollment with an odds ratio (OR) of 0.25 (95% CI, 0.06-0.74) and less in-hospital length of stay, compared to non-metformin group. There were no statistical differences between the metformin group and non-metformin group for all three inflammatory markers (ferritin, CRP, and IL-6) on admission or peak levels during hospitalization. Furthermore, meta-analysis demonstrated that metformin use was significantly correlated with reduced mortality with an OR of 0.73 (95% CI, 0.65-0.82) in the fixed effect model and 0.58 (95% CI, 0.42-0.80) for the random effect model.

**Conclusions:** Our results provide supporting evidence that metformin may confer increased survival in patients with COVID-19 and T2DM treated with Metformin prior to hospitalization.

## Introduction

Rapid outbreak and spread of coronavirus disease 2019 (COVID-19), caused by the novel severe acute respiratory syndrome coronavirus 2 (SARS-CoV-2), leads to a global health crisis ^1-5^. Risk factors and comorbidities linked to worse outcomes for COVID-19 patients have been identified. These include old age ^5^, chronic pulmonary disease and smoking ^6^, cardiovascular disease ^6^, hypertension ^7^, diabetes mellitus and obesity ^8^. Patients with pre-existing diabetes mellitus, depending on nations and patient cohorts, have been reported to account for patients with COVID-19 from 5-20% in China, 17% in Lombardy in Italy to 33% in the US ^1-5^. Diabetic patients hospitalized with COVID-19 are two- to three-fold more likely to be admitted into intensive care units than that of non-diabetics and the mortality rate is at least doubled ^6,9-11^, suggesting urgent need for effective treatments in patients with COVID-19 and diabetes mellitus.

Metformin is widely used as the first-line therapy for type 2 diabetes mellitus (T2DM) ^12^. Metformin is effective, safe, and inexpensive and may reduce the risk for cardiovascular events and death in T2DM ^13^. Historically, because of its host-directed anti-viral properties, metformin was used during the treatment of influenza outbreak ^14^. Likewise, based on the pathogenesis of SARS-CoV-2, several mechanisms have been speculated about the possible beneficial effects of metformin in COVID-19 patients with pre-existing T2DM, such as anti-inflammatory effects ^15^, reduction in neutrophils ^16^, increasing the cellular pH to inhibit viral infection, interfering with the endocytic cycle and reversing established lung fibrosis ^17^. In contrast, metformin use has also been challenged in COVID-19 patients with T2DM because of the potential risk for lactic acidosis, particularly in the cases of multi-organ failure and for promoting SARS-CoV-2 infection by possibly increasing in ACE2 availability in the respiratory tract ^18^. Recent accumulating evidence suggests that treating COVID-19 patients with pre-existing T2DM with metformin may not be harmful, but whether metformin endows a protective effect and offers a low mortality in patients with COVID-19 and T2DM remains inconclusive. Therefore, more clinical data regarding the use of metformin in COVID-19 patients with pre-existing T2DM is needed to provide clinical evidence. In the present study, we aimed to address whether metformin use is beneficial in COVID-19 patients with pre-existing T2DM. We found that COVID-19 patients with pre-existing T2DM treated with metformin prior to hospitalization had favorable outcome compared to metformin non-users. Our findings support the use of metformin as the first-line therapy for T2DM in patients with COVID-19 prior to hospitalization.

## Materials and methods

### Study Design and Participants

We conducted a retrospective study of COVID-19 patients admitted to the St Luke’s University Health Network (SLUHN) from 3/16/2020 to 2/15/2021. COVID-19 is defined by positive results of RT-PCR for COVID-19 nasal swab specimens. The inclusion criteria included patients (Age ≥ 18) with COVID-19 and pre-existing T2DM. The exclusion criteria were incomplete medical records, lack of outcome data, type 1 DM, and in-hospital length of stay less than 1 day. Oral hypoglycemic medications including metformin were held during hospitalization according to the SLUHN policy. Patients with pre-existing T2DM who took metformin prior to admission were designated as the metformin group and those did not receive metformin prior to admission were assigned as the non-metformin group. Metformin use was determined as documented in the home medication list in EPIC within the 3 months before admission. The primary outcome was a composite of in-hospital death or hospice care enrollment. The secondary outcomes were in-hospital length of stay and mechanical ventilation. This study was approved by the SLUHN institutional review board (SLIR 2020-100) and was granted a waiver of informed consent.

### Multivariate regression

To investigate the potential effect of metformin on the primary outcome, multivariate regression was applied. Clinically relevant confounders including age, height, weight, gender, race, comorbidities (congestive heart failure (CHF), coronary heart disease (CAD), asthma, chronic obstructive pulmonary disease (COPD)), clinical lab tests (BUN, creatinine, ALT, total bilirubin) were entered into a multivariate logistic regression model as covariates. To avoid the introduction of bias, no imputations were performed for missing values. Patients with missing values were excluded from the analysis.

### Propensity score matching (PSM) and inverse probability of treatment weighting (IPTW)

To account for the potential confounders associated with the use of metformin and to ensure the robustness of our results, propensity score matching was used based on variables including age, height, weight, gender, race, comorbidities (CHF, CAD, asthma, COPD), clinical lab tests (BUN, creatinine, ALT, total bilirubin). Propensity scores for each patient were estimated by a multivariate logistic regression model. The matched cohort was created at a 1:1 ratio with a caliper size of 0.05. Using the estimated propensity scores as weights, the IPTW method was used to generate additional weighted cohort (IPTW cohort). The balance between covariates was evaluated by estimating standardized mean differences (SMD) in both the propensity score matched cohort and the IPTW cohort.

### Meta-analysis

To assess whether metformin use prior to admission offers a low mortality by comparing and integrating the results of different studies, meta-analysis was carried out as described previously^19^. Odds ratio (OR) was used as the summary statistics. The OR represents the odds of a death event occurring in the metformin group in comparison to the non-metformin group. The point of estimate of the OR is considered statistically significant at the P < 0.05 level if the 95% confidence interval does not include the value 1. In our study, fixed and random effect models were employed. Studies with metformin treatment prior to admission in patients with COVID-19 and pre-existing T2DM were included.

### Statistical analysis

Data were presented as mean and 95% confidence interval (CI) or median and interquartile range (IQR) for continuous variables. For differences in continuous variables, Student’s t tests (normally distributed) or Mann-Whitney U test (non-normally distributed) were used for comparison. Categorical variables were expressed as percentage (%). For the comparison of categorical variables, χ2 tests were used. The risk of primary outcome was calculated as OR using multivariate logistic regression models. P values < 0.05 were considered to be statistically significant. All analyses were conducted with R (version 3.6.1).

## Results

### Baseline characteristics

A total of 1872 admissions with confirmed COVID-19 and diabetes from 11 hospitals in the St Luke’s University Health Network were initially included for this study. After excluding multiple admissions (hospital length of stay less than 24 hours, type 1 diabetes, or missing value), we included 1356 patients in our study cohort (Fig. 1). In the final cohort, there were 361 patients taking metformin prior to admission and 995 patients treated with anti-diabetic other than metformin. The baseline characteristics of our cohort on admission are summarized in Table 1.

**Figure 1.**
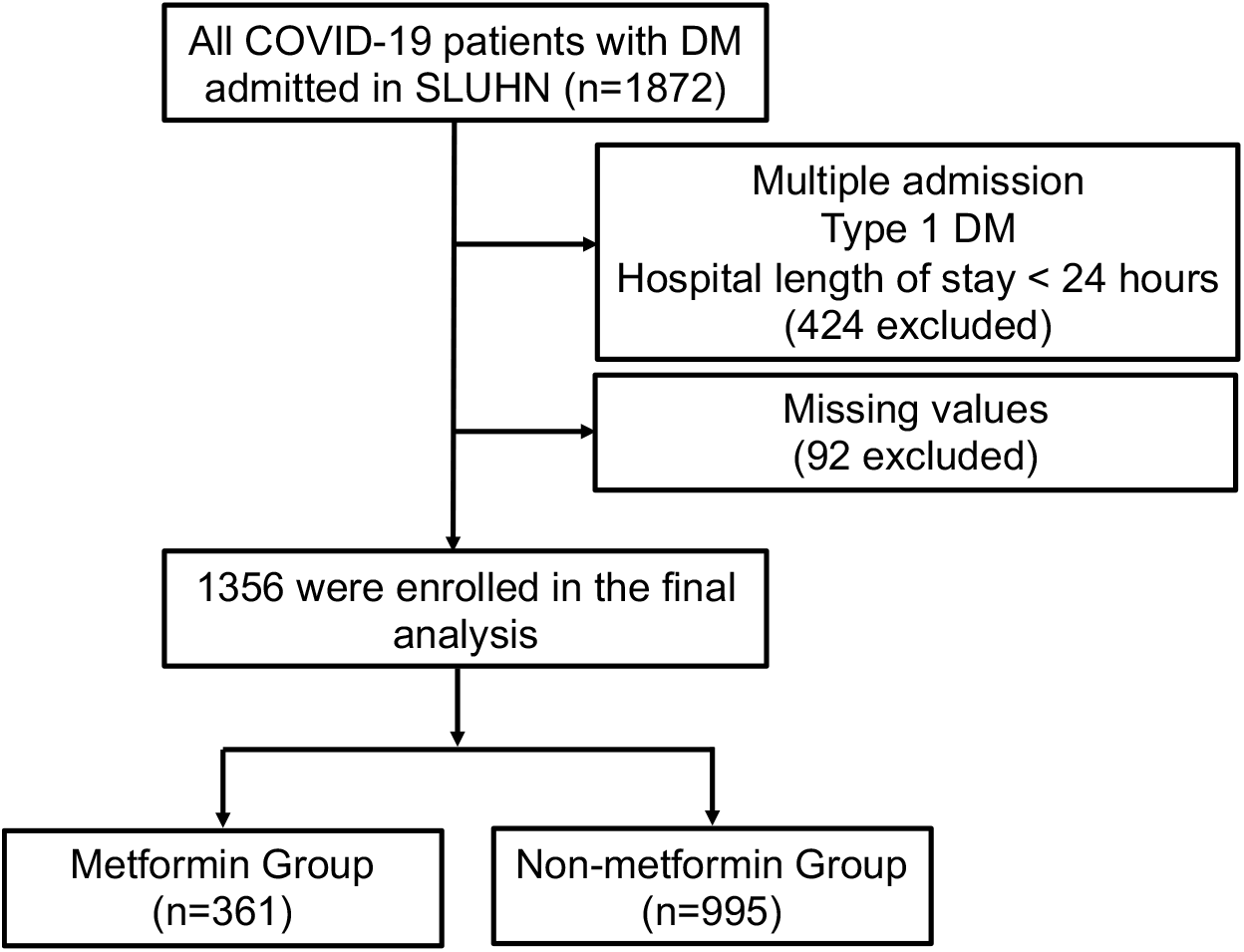
Flowchart illustration of the study cohort.

**Table 1.** Comparisons of demographic and clinical characteristics between the original cohort and matched cohort.

| Covariate | Original cohort |  |  | Matched cohort |  |  |
| --- | --- | --- | --- | --- | --- | --- |
|  | Non-metformin | Metformin | SMD | Non-metformin | Metformin | SMD |
| N | 995 | 361 |  | 360 | 360 |  |
| Age |  |  | 0.262 |  |  | 0.075 |
| <50 | 92 | 45 |  | 46 | 44 |  |
| 50-80 | 692 | 274 |  | 255 | 274 |  |
| >80 | 211 | 42 |  | 59 | 42 |  |
| Male (%) | 538 (54.1) | 218 (60.4) | 0.128 | 218 (60.6) | 217 (60.3) |  |
| Weight-kg |  |  | 0.142 |  |  | 0.04 |
| Median | 89.60 | 92.52 |  | 91.40 | 92.50 |  |
| Interquartile range | 74.73-107.99 | 78.60-111.11 |  | 76.71-110.97 | 78.57-111.00 |  |
| Height-m |  |  | 0.178 |  |  | 0.034 |
| Median | 167.64 | 170.2 |  | 170.2 | 170.2 |  |
| Interquartile range | 160.02-177.16 | 162.6-177.8 |  | 162.6-177.9 | 162.6-177.8 |  |
| Race |  |  | 0.068 |  |  | 0.055 |
| Asian | 14 (1.4) | 7 (1.9) |  | 7 (1.9) | 7 (1.9) |  |
| Black | 105 (10.6) | 44 (12.2) |  | 38 (10.6) | 44 (12.2) |  |
| White | 737 (74.1) | 262 (72.6) |  | 265 (73.6) | 262 (72.8) |  |
| Unknown | 139 (14.0) | 48 (13.3) |  | 50 (13.9) | 47 (13.1) |  |
| Comorbidity-no. (%) |  |  |  |  |  |  |
| Hypertension | 846 (85.0) | 287 (79.5) | 0.145 | 288 (80.0) | 286 (79.4) | 0.014 |
| CHF | 330 (33.2) | 65 (18.0) | 0.353 | 54 (15.0) | 65 (18.1) | 0.082 |
| CAD | 298 (29.9) | 77 (21.3) | 0.198 | 71 (19.7) | 77 (21.4) | 0.041 |
| COPD | 205 (20.6) | 51 (14.1) | 0.172 | 47 (13.1) | 51 (14.2) | 0.032 |
| Asthma | 127 (12.8) | 58 (16.1) | 0.094 | 56 (15.6) | 58 (16.1) | 0.015 |
| Laboratory tests |  |  |  |  |  |  |
| ALT | 40.33 (35.70) | 48.07 (37.63) | 0.211 | 42.29 (32.09) | 48.03 (37.68) | 0.164 |
| BUN | 30.16 (22.07) | 22.28 (14.67) | 0.421 | 21.33 (13.40) | 22.32 (14.66) | 0.071 |
| Creatinine | 1.89 (1.98) | 1.27 (0.57) | 0.428 | 1.27 (0.65) | 1.27 (0.57) | 0.005 |
| Total Bilirubin | 0.70 (2.77) | 0.61 (0.35) | 0.047 | 0.62 (0.42) | 0.61 (0.35) | 0.016 |

CHF, congestive heart failure; CAD, coronary artery disease; COPD, chronic obstructive pulmonary disease; ALT, alanine aminotransferase; BUN, blood urea nitrogen

### Primary outcomes

The primary outcome was a composite of in-hospital death or hospice. Of the 361 in the metformin group, primary outcome events occurred in 3 patients (0.83%) including 2 of in-hospital death and 1 for hospice, compared with 40 of 995 (4.02%) in the non-metformin group, specifically 21 of in-hospital death and 19 for hospice. In the multivariate logistic regression analyses, after adjusting age, gender, race, comorbidities, BUN, creatinine, ALT, total bilirubin, outpatient metformin use was significantly associated with lower primary outcome events (OR, 0.25; 95% CI, 0.06-0.74; P = 0.027) compared to non-metformin (Fig. 2).

**Figure 2.**
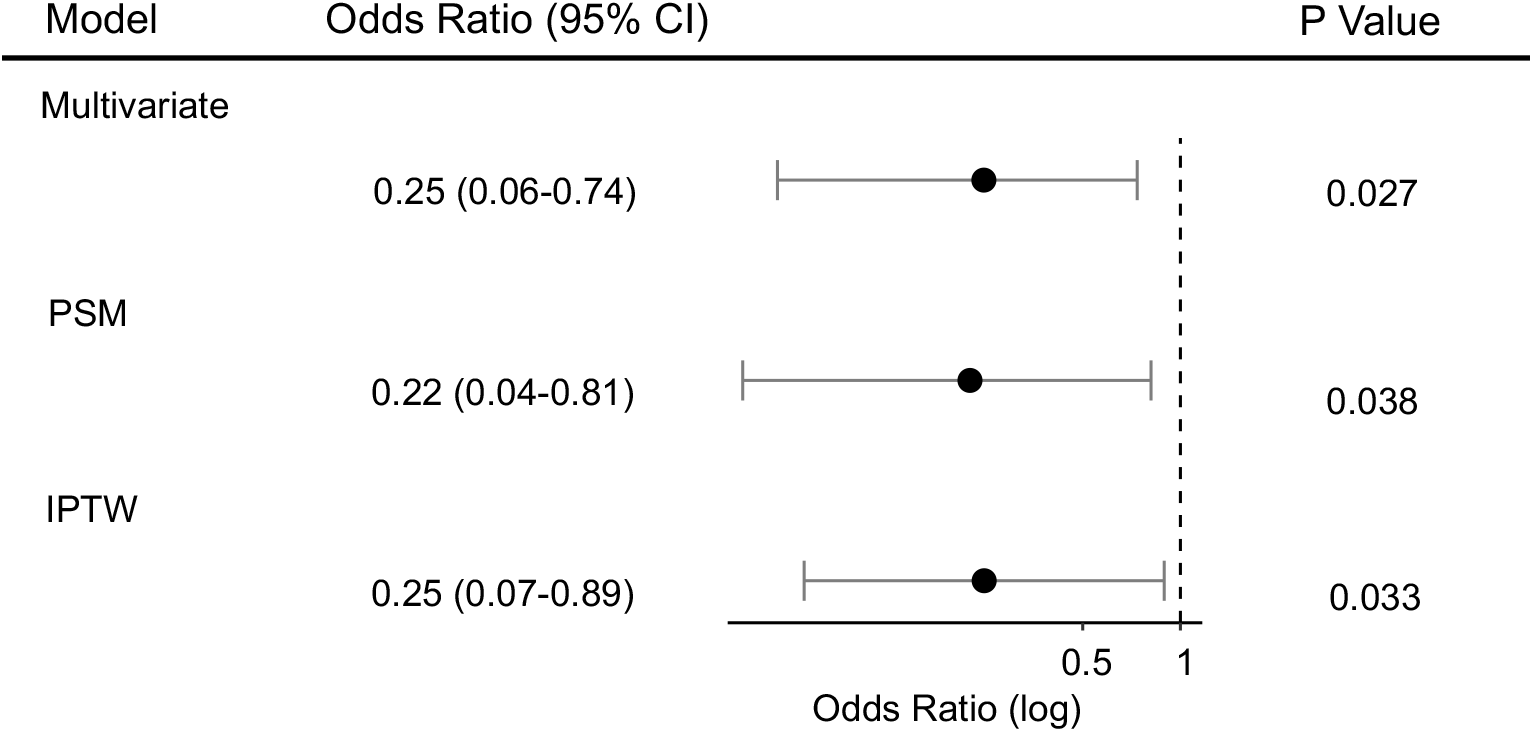
Primary outcome analysis with three different models. Outpatient metformin use was significantly associated with reduced primary outcomes. PSM, propensity score matching; IPTW, inverse probability of treatment weighting.

To verify the robustness of the model, we conducted a sensitivity study with randomly excluding patients from two hospitals. The adjust OR from this new cohort (n=1143) for the primary outcome was 0.25 (95% CI, 0.06-0.76; P = 0.031). To account for confounding that could lead to the protective association between metformin and the primary outcome, we further performed propensity score matching (PSM) and propensity score-based inverse probability of treatment weighting (IPTW) analyses. After PSM or IPTW, the covariates between the metformin group and the non-metformin group were well balanced based on SMD as shown in Table 1 and Fig. 3. In the PSM matched cohort, after adjusting the above variables, there was a reduced risk of primary outcome events in the metformin group (adjusted OR, 0.22; 95% CI, 0.04-0.81; P = 0.038) (Fig. 2). Similarly, in the IPTW matched cohort, the adjusted OR for the primary outcome was 0.25 (95% CI, 0.07-0.89; P = 0.033).

**Figure 3.**
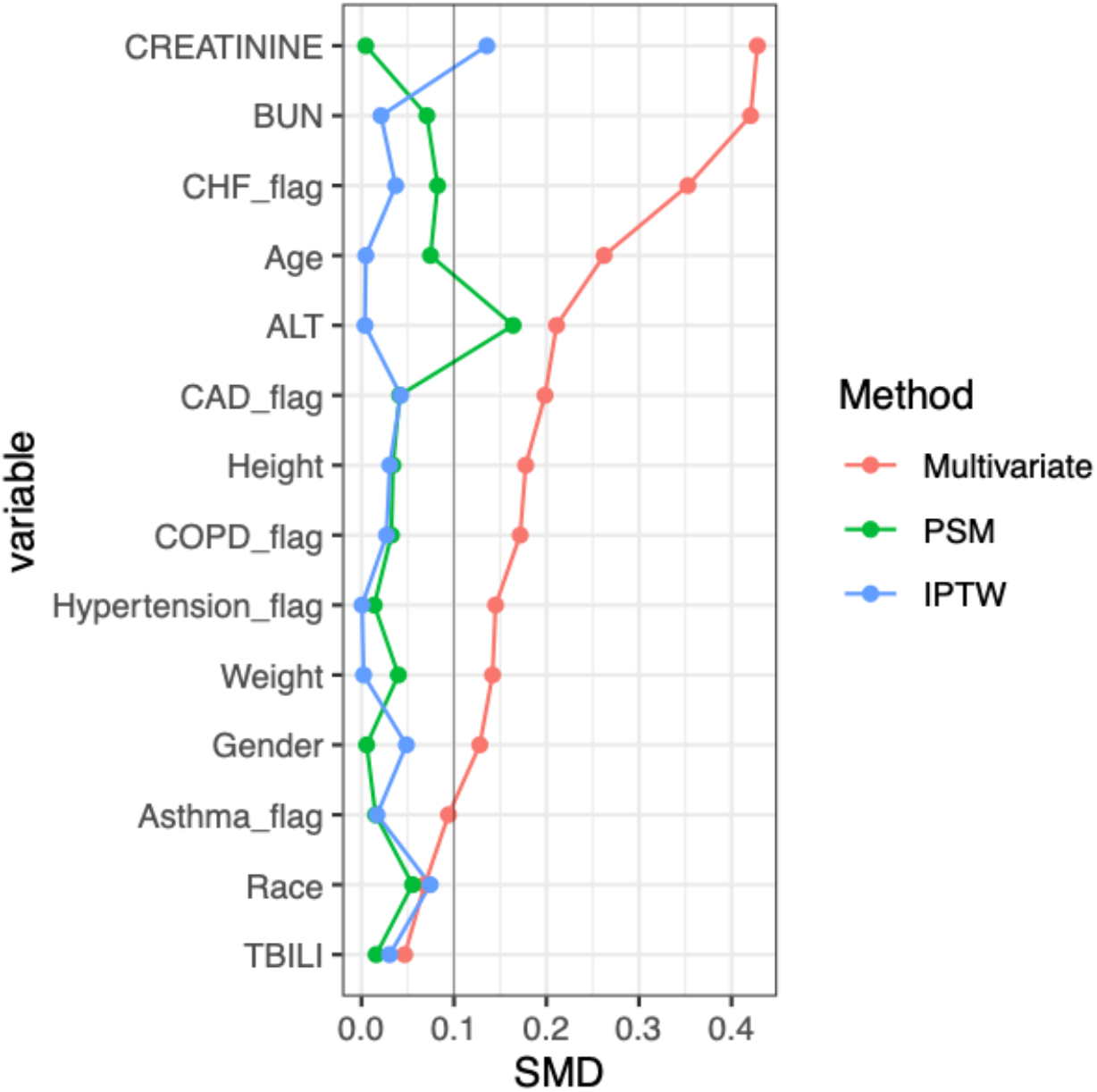
Change in standardized mean difference (SMD) before and after matching. PSM, propensity score matching; IPTW, inverse probability of treatment weighting; CHF, congestive heart failure; CAD, coronary artery disease; COPD, chronic obstructive pulmonary disease; ALT, alanine aminotransferase; BUN, blood urea nitrogen; TBILI, total bilirubin.

### Secondary outcomes

To further investigate the potential effects of metformin use prior to admission, we accessed in-hospital length of stay and the rate of mechanical ventilation in the original cohort and PS matched cohort (Table 2). The in-hospital length of stay was significantly lower with a mean of 6.42 ± 5.51 days in the metformin group compared to 8.04 ± 7.78 days in the non-metformin group (P < 0.001) in the original cohort. A similar result was maintained (P = 0.049) in the propensity score matched cohort with a mean of 6.43 ± 5.51days in the metformin group and 7.38 ± 7.40 days in the non-metformin group, respectively. For mechanical ventilation, while the rate (3.3%) in the metformin group was markedly smaller than that (6.7%) in the non-metformin group (P = 0.018) in the original cohort, there was no difference in the matched cohort.

**Table 2.**
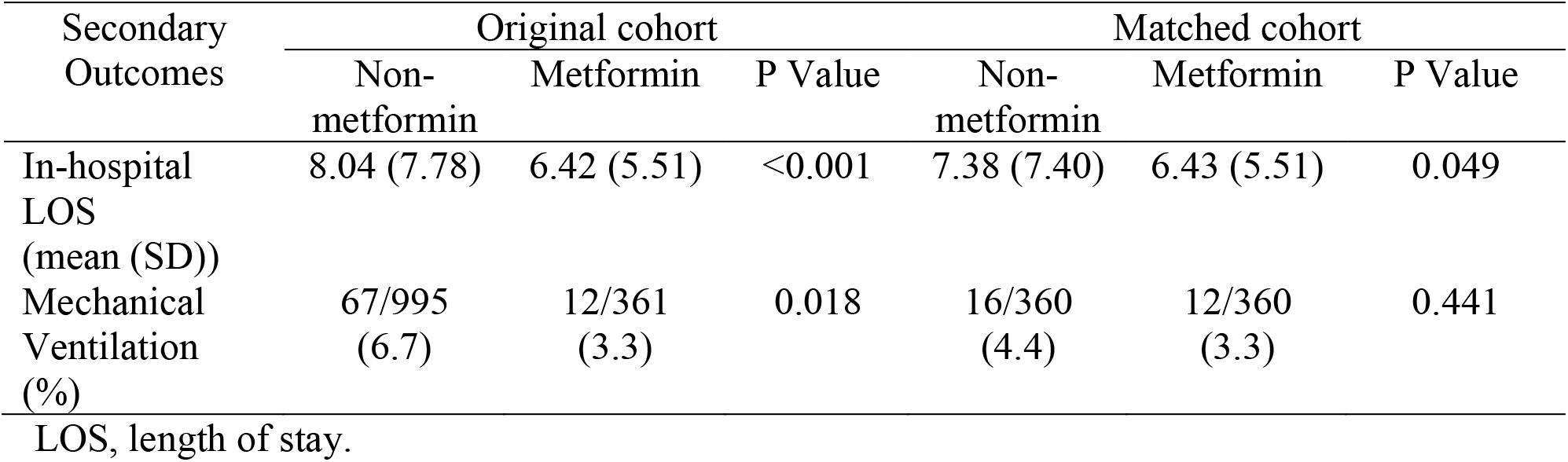
Secondary outcome analysis of the original cohort and matched cohort

| Secondary Outcomes | Original cohort |  |  | Matched cohort |  |  |
| --- | --- | --- | --- | --- | --- | --- |
|  | Non-metformin | Metformin | P Value | Non-metformin | Metformin | P Value |
| In-hospital LOS (mean (SD)) | 8.04 (7.78) | 6.42 (5.51) | <0.001 | 7.38 (7.40) | 6.43 (5.51) | 0.049 |
| Mechanical Ventilation (%) | 67/995 (6.7) | 12/361 (3.3) | 0.018 | 16/360 (4.4) | 12/360 (3.3) | 0.441 |

LOS, length of stay.

### Inflammatory responses in the metformin group versus the non-metformin group

Metformin has been shown to have anti-inflammatory effects ^15^. The inflammatory response has been shown to play a pivotal role in the progression of COVID-19, and inflammatory cytokine storm ultimately results in severe COVID-19. To determine whether use of metformin prior to admission affects inflammatory responses in COVID-19 patients, we compared the measured serum levels of ferritin, C-reactive protein (CRP) and interlukin-6 (IL-6) on admission and peak levels during hospitalization between the metformin group and the non-metformin group. In this retrospective cohort, serum ferritin was measured in 92.2 % of cases, CRP was measured in 91.5 % of cases and IL-6 was measured in 39.0% of cases. There were no statistical differences between the metformin group and non-metformin group for all three markers on admission or peak levels during hospitalization (Fig. 4).

**Figure 4.**
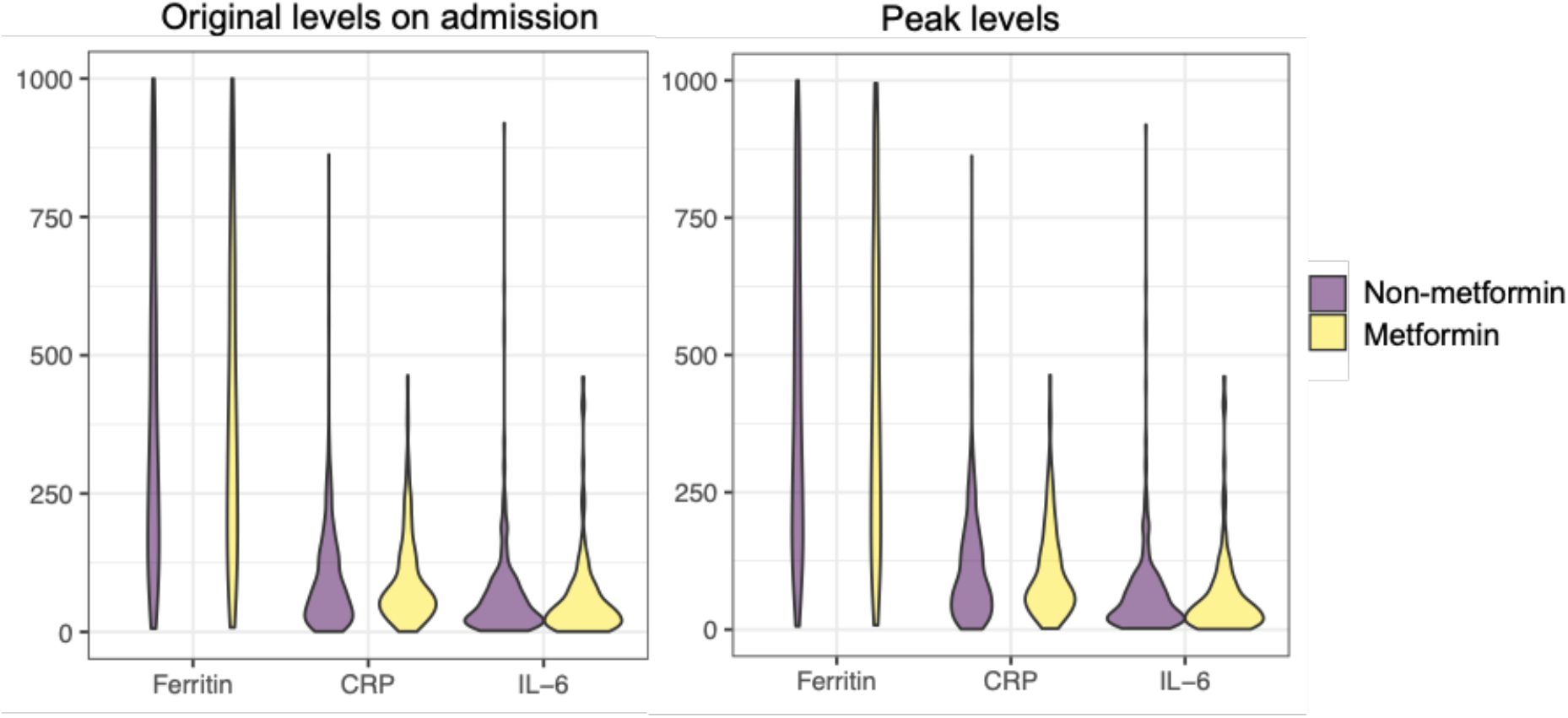
Measurement of serum levels of ferritin, CRP and IL-6 on admission and peak values during hospitalization between the metformin group and non-metformin group. Violin plot data were derived from the original cohort. The units for ferritin, CRP and IL-6 were ng/mL, mg/L and pg/mL, respectively.

### Meta-analysis of in-hospital mortality in the metformin group versus the non-metformin group

A few of retrospective observational studies in COVID-19 patients with T2DM suggested that metformin treatment prior to admission is associated with a potential reduction in in-hospital mortality ^20-24^. To further access the association between metformin use and decreased mortality by increasing statistical power, we performed a meta-analysis of 6 studies with the use of metformin before admission including the present study to determine the effects of metformin on mortality. Despite the heterogeneity among the studies, the pooled OR was 0.73 (95% CI, 0.65-0.82) in the fixed effect model and 0.58 (95% CI, 0.42-0.80) in the random effect model, indicating metformin use was associated with a significant reduction of in-hospital death (Fig. 5).

**Figure 5.**
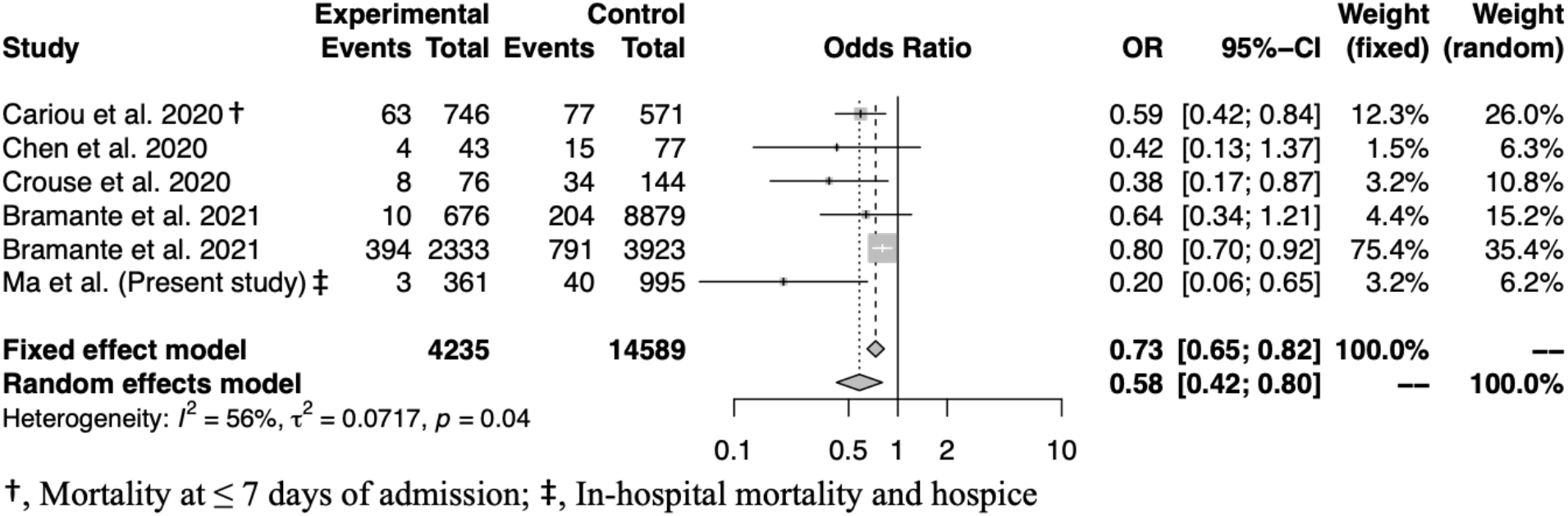
Meta-analysis of selected studies with the present study comparing in-hospital mortality between outpatient metformin use and non-metformin use in patients with COVID-19 and pre-existing T2DM.

## Discussion

With the pandemic of COVID-19 and its high mortality especially in patients with pre-existing diabetes mellitus, there is an urgent need to develop treatments that are effective, safe, and available immediately. Metformin is a widely used anti-diabetic agent that is safe, effective, and inexpensive for T2DM. In view of host-directed anti-viral properties and anti-inflammatory effects of metformin, we set out to explore whether metformin is beneficial in patients with COVID-19 and preexisting T2DM. In this retrospective cohort study, we found that metformin therapy prior to admission in patients with COVID-19 and preexisting T2DM was significantly associated with reduced primary outcome events, which was a composite of in-hospital mortality and enrollment in hospice care. Of note, most patients enrolled in hospice/comfort care passed away shortly in the setting of COVID-19 infection in our cohort. In agreement with our findings, in CORONADO study, a large observational multicenter French study, metformin users prior to hospital admission were associated with a significant reduction in mortality at day 7, compared to non-users in an unadjusted analysis (OR: 0.59, 95% CI: 0.42–0.84). There was a trend but not significant decrease in mortality after the full adjustment ^22^. In a retrospective study of 120 Chinese patients with pre-existing diabetes and confirmed and unconfirmed but clinically diagnosed COVID-19, there was a reduced but not statically significant trend for in-hospital mortality in metformin users (n = 43), compared to metformin non-users (9.3% vs 19.5%) ^23^. Consistent with studies in China and French, a recent retrospective study in USA with 220 COVID-19 patients with pre-existing diabetes including 76 metformin users and 144 non-users showed that metformin treatment was independently and significantly associated with a reduction in mortality in COVID-19 with T2DM (OR 0.33; 95%CI 0.13-0.84; p=0.0210) ^24^. In another large US retrospective study of 6,256 subjects (>95% diabetes, 52.8% female, mean age 75 years) from UnitedHealth database, a significantly decreased mortality in women (but not in men) was observed in metformin users with an odds ratio of 0.759 by propensity matching and an hazard ratio of 0.785 by Cox proportional-hazards, compared to metformin non-users ^21^. Furthermore, retrospective cohort analysis of US electronic health record (EHR) data from 12 hospitals and 60 primary care clinics in the Midwest have shown that outpatient metformin use was associated with a decrease in mortality from COVID-19, OR 0.32 (P =0.002), and a trend towards decreased admission for COVID-19 ^20^. These studies indicate the possible benefit of metformin therapy prior to admission in COVID-19 patients with pre-existing T2DM. Furthermore, in view of small number of patients in most of above observational studies and lack of statistical power, meta-analysis was carried out in the present study to integrate the results of different studies including our study. We found that metformin use prior to admission was associated with a significant reduction of in-hospital death by 27% with a pooled OR of 0.75 (95% CI, 0.66-0.84) in the fixed effect model and by 42% with a pooled OR of 0.58 (95% CI, 0.42-0.80) in the random effect model.

While metformin therapy prior to admission is significantly associated with reduced mortality, there is evidence suggesting metformin treatment might be effective in patients with COVID-19 and pre-existing T2DM during hospitalization. In a retrospective analysis in China of 283 COVID-19 patients with pre-existing T2DM, treating patients with metformin (n = 104) was associated with a significantly lower in-hospital mortality, compared with metformin non-users (n = 179) with similar patients characteristics, laboratory parameters, and treatments (2.9% vs 12.3%, respectively; P = 0.01), despite significantly higher fasting blood glucose levels in metformin users^25^. Furthermore, another retrospective study of 1213 hospitalized COVID-19 patients with pre-existing T2DM on the use of metformin from 16 hospitals in China found that metformin was not only associated with an increased incidence of acidosis (adjusted hazard ratio 2.45, 95% CI 1.08-5.54, P = 0.032), but with a significant reduction in heart failure (adjusted hazard ratio 0.61, 95% CI 0.43-0.87, P = 0.006) as well as inflammatory responses, although there were no differences in 28-day COVID-19-related mortality or the length of hospitalization ^26^. These results further support continuing use of metformin in COVID-19 patients with pre-existing T2DM with closely monitoring acidosis and kidney function.

In response to SARS-CoV-2 infection, it seems that there are two distinct but overlapping pathologic phases; the first viral response phase triggered by the virus itself and the second inflammatory phase medicated by the host response ^27^. Accumulating evidence indicates that many patients with severe COVID-19 manifest cytokine storm syndrome. Therefore, agents with evidence of reducing hyper-inflammation in COVID-19 patients could be therapeutic. It has been reported that metformin has anti-inflammatory effects ^15^ and can modulate the immune responses and restore immune homeostasis in immune cells via AMPK-dependent mechanisms ^28^. In a retrospective study of 120 Chinese patients with pre-existing diabetes and confirmed and unconfirmed but clinically diagnosed COVID-19, a significantly lower increase in interleukin-6 was observed in metformin users prior to admission, compare to non-users (n = 77) ^23^. In addition, the dynamic trajectories of serum inflammatory factors, including CRP, IL-6, IL-2, and tumor necrosis factor-alpha (TNF-α), have demonstrated lower degrees of elevation in the hospitalized COVID-19 patients with metformin treatment than the non-metformin group, particularly in patients with severe COVID-19 ^26^. Conversely, there were no significant differences in the levels of ferritin, CRP, or IL-6 on admission and peak value during hepatization between metformin users and non-metformin users in our study. More studies are needed to clarify whether metformin could mitigate inflammatory responses in patients with COVID-19.

Several limitations are present in our study. First, although efforts were made to balance and control for potential confounding factors by multiple variable adjustments and propensity score matching, due to the inherent nature of retrospective observational studies, residual confounders are likely to exist and could not be balanced. Second, a positive effect was observed in the meta-analysis with an overall reduction of mortality, but the size of the retrospective studies that were included in the meta-analysis varied considerably, resulting in moderate-to-high heterogeneity (I^2^ = 56%, P=0.04). Future prospective randomized controlled trials are needed to determine any definitive clinical beneficial outcomes in patients with COVID-19 and pre-existing T2DM treated with metformin.

In conclusion, our findings demonstrate that metformin therapy prior to admission in patients with COVID-19 and pre-existing T2DM is associated with a significant reduction of in-hospital mortality and provide clinical evidence in support of metformin as first line anti-diabetic agent for patients with COVID-19 and pre-existing T2DM.

## Data Availability

De-identified data is available from the corresponding author on reasonable request

## Author Contributions

ZM and MK designed the study and wrote the manuscript. ZM, NP, and PV collected the data. ZM performed and analyzed data. All authors reviewed the results and approved the final version of the manuscript.

## Acknowledgements

We thank Dr. Douglas S. Corwin for helpful discussion with data collection and interpretation.

